# Multilevel Interventions to Improve Colonoscopy Completion After Abnormal Fecal Immunochemical Testing in a Safety-Net Health System: Preliminary Findings from the IMPACTT Study

**DOI:** 10.64898/2026.11.10.26367784

**Authors:** Urmimala Sarkar, Jeanette Wong, Kristan Olazo, Sara Colom Brana, Shreya Patel, Rena Pasick, Charles E. McCulloch, Ma Somsouk

## Abstract

**Background:** Timely diagnostic colonoscopy is a critical step after an abnormal fecal immunochemical test (FIT), yet many patients do not complete follow-up within recommended timeframes, particularly in safety-net settings. Barriers occur across multiple levels and stages of care, suggesting a need for interventions that address both clinic processes and patient-level barriers.

**Methods:** IMProving Adherence to Colonoscopy through Teams and Technology (IMPACTT) evaluated two concurrent interventions across 12 adult primary care clinics in the San Francisco Health Network. A clinic-level Best Practice Bundle (BPB), implemented in a stepped-wedge rollout, included patient tracking, standardized electronic health record documentation, standardized education and communication, and audit and feedback. In parallel, we randomly assigned eligible patients to a patient-level instruction and navigation (PIN) intervention or usual care. PIN provided targeted outreach to patients who were neither referred nor scheduled, as well as multilingual text reminders that allowed patients to confirm, cancel, or reschedule their colonoscopy. The primary outcome was colonoscopy completion within 180 days of an abnormal FIT. This preliminary analysis reports unadjusted comparisons among patients with sufficient follow-up at the data cutoff.

**Results:** Among 2,303 patients with an abnormal FIT, 1,136 were identified before and 1,167 after their clinic implemented the BPB. Colonoscopy completion within 180 days was 44.1% before BPB implementation and 60.1% after implementation (risk difference, 16.0 percentage points; 95% CI, 11.8 to 20.1; p<0.001). Referral to gastroenterology within 42 days increased from 69.8% to 84.2%, and among patients who completed colonoscopy, the median time from FIT to colonoscopy decreased from 131 to 86 days. In the randomized PIN trial, we assigned 1,415 patients to usual care (n=698) or PIN (n=717). Colonoscopy completion within 180 days did not differ between usual care and PIN (58.4% vs 57.7%; risk difference, -0.7 percentage points; 95% CI, -5.9 to 4.6; p=0.9). PIN also did not improve referral within 42 days or scheduling within 56 days.

**Conclusions:** In preliminary unadjusted analyses, implementation of a clinic-level bundle targeting tracking, communication, documentation, and feedback was associated with improved timely follow-up after abnormal FIT, whereas a randomized technology-supported patient instruction and navigation intervention did not improve colonoscopy completion. Prespecified analyses accounting for calendar time, clinic-level clustering, patient characteristics, and concurrent intervention assignment are forthcoming.

**Trial registration:** ClinicalTrials.gov NCT06191185.

## Introduction

Colorectal cancer (CRC) screening reduces CRC incidence and mortality, yet disparities persist among racial and ethnic minority and socioeconomically vulnerable populations.^1,2^ Fecal immunochemical testing (FIT) is scalable and is preferred by many patients, including those served by safety-net health systems.^1,3^ However, the benefit of FIT screening depends on completing diagnostic colonoscopy after an abnormal result.^4^ Delays in or failure to complete colonoscopy after an abnormal FIT are associated with increased risk of CRC, advanced-stage disease, and CRC mortality.^5–7^

Completing colonoscopy after an abnormal FIT requires successful transitions across multiple steps and levels of care, including communicating the abnormal result, referral from primary care, scheduling by gastroenterology, and patient preparation and attendance.^8,9^ In the San Francisco Health Network (SFHN), prior work found that only 52% of patients with an abnormal FIT completed colonoscopy within one year, with losses occurring at referral, scheduling, and procedure attendance.^9^ Barriers span the health system, care team, and patient levels, including inadequate tracking, lack of standardized team workflows, comorbid conditions, transportation and social support needs, housing instability, and language and communication barriers.^9,10^ These findings suggest that improving follow-up requires interventions that address both clinic processes and patient-level barriers.

A systematic review identified patient navigation, provider reminders, and provider performance data as promising strategies to improve colonoscopy follow-up, while highlighting limitations in the quality and consistency of the available evidence.^4^ However, results have been inconsistent, and multicomponent, multilevel approaches have not been comprehensively evaluated in resource-constrained safety-net settings.^4,8^ We therefore conducted IMProving Adherence to Colonoscopy through Teams and Technology (IMPACTT), a multilevel study implemented across 12 primary care clinics in an urban safety-net health system. IMPACTT evaluated a clinic-level Best Practice Bundle (BPB) implemented in a nonrandomized stepped-wedge rollout alongside a randomized patient-level instruction and navigation (PIN) intervention. We report preliminary unadjusted findings for colonoscopy completion and intermediate steps in diagnostic follow-up.

## Methods

### Study design and setting

IMProving Adherence to Colonoscopy through Teams and Technology (IMPACTT; NCT06191185) tested a multilevel intervention to improve colonoscopy after an abnormal fecal immunochemical test (FIT). The protocol has been published.^11^ The trial was conducted in all 12 adult primary care clinics of the San Francisco Health Network (SFHN), a publicly funded, urban safety-net system. All 12 clinics refer patients to a single gastroenterology (GI) unit at Zuckerberg San Francisco General Hospital and share one electronic health record (EHR) (Epic Systems, Verona, WI).

We implemented two interventions concurrently. We introduced a clinic-level Best Practice Bundle (BPB) in a stepped-wedge rollout. We randomly assigned individual patients to the patient-level instruction and navigation (PIN) intervention or usual care throughout the study, before, during, and after each clinic received the BPB, so that the effects of each intervention could be estimated independently. We registered the trial as a 2 × 2 factorial design (BPB × PIN) on December 20, 2023, before enrollment began on July 8, 2024; data were analyzed through September 15, 2026. The UCSF Institutional Review Board approved the study with a waiver of informed consent. We used the CONSORT statement to guide reporting of the randomized PIN component and applicable STROBE recommendations to guide reporting of the nonrandomized clinic-level BPB component.

### Participants

We included all SFHN patients aged 45 to 75 years with an abnormal FIT result empaneled at one of the 12 clinics. Patients were not excluded by language. We identified eligible patients through a weekly EHR query and entered them into a registry in the UCSF Research Analysis Environment, a secure research platform.

### Clinic-level intervention: Best Practice Bundle

The BPB comprised four strategies:

1. Patient tracking: We trained staff to run an EHR report listing patients with a referral or scheduling care gap.
2. Standardized EHR documentation: telephone encounters, entry of abnormal FIT (ICD-10 R19.5) on the problem list, standard phrases, direct-access colonoscopy referral, and team routing.
3. Standardized education and communication: dot-phrases, key messages, and health-coaching scripts on the meaning of an abnormal FIT, scheduling the procedure, and steps to colonoscopy.
4. Audit and feedback: We provided a report of each clinic’s care gap every 2 months and a quarterly implementation survey.

The study team introduced the BPB during a meeting with clinic leadership and provided materials and follow-up by email. One or two clinics began the BPB each quarter over 2 years. The order of clinic entry followed clinic leadership’s preferred timing and was not randomized. We classified each patient as belonging to the pre-BPB or BPB period based on whether the patient’s clinic had started the BPB at the time of the abnormal FIT.

### Patient-level intervention: PIN

We randomly assigned patients to PIN or usual care. PIN assignment prompted action at each step of the care continuum:

1. Unreferred: If no GI referral was placed within 21 days of the abnormal FIT, the study team sent the primary care provider (PCP) a secure EHR message.
2. Unscheduled: If colonoscopy was not scheduled within 14 days of referral, the team notified the PCP and then called the patient to explain the result and give the GI scheduling number.
3. Scheduled: Patients scheduled for colonoscopy received a text reminder in English, Spanish, or Chinese 7 days before the procedure (HealthySMS, Audacious Software, Chicago, IL) that allowed them to confirm, cancel, or reschedule by numeric response. Patients who did not respond received a follow-up call from study staff 2 days before the procedure.

Patients assigned to usual care received the clinic’s standard referral and scheduling processes, including the BPB once BPB was implemented at the clinic. For ethical reasons, the study team also messaged the PCP of any patient in either arm without a GI referral at 180 days.

### Outcomes

The primary outcome was completion of colonoscopy within 180 days of the abnormal FIT. Secondary outcomes were GI referral within 42 days, colonoscopy scheduled within 56 days, and days from FIT to referral, scheduling, and colonoscopy. Process measures for the PIN arm were text-message response and disposition (confirmed, canceled, rescheduled). We ascertained all outcomes from the EHR. We compared scheduling within 56 days only between PIN arms because scheduling dates were not accurately captured before the BPB.

### Statistical analysis

We summarized baseline characteristics as median (interquartile range, IQR) or number (%) and compared groups with the Wilcoxon rank-sum test for continuous variables and the chi-square or Fisher exact test for categorical variables. We report each binary outcome as a proportion with a 95% confidence interval (CI) and between-group differences as risk differences with 95% CIs. We report days to each step as median (IQR) among patients who reached that step and, in sensitivity analyses, capped at 180 days.

We analyzed the 180-day outcome among patients whose 180-day window had closed by the data cutoff (September 15, 2026). We analyzed referral at 42 days and scheduling at 56 days among patients whose respective follow-up windows had closed. As prespecified, we counted patients who left the network before 180 days as not completing colonoscopy.

This is a preliminary report of unadjusted comparisons; follow-up continues to the registered primary completion date of November 27, 2026. The prespecified mixed-effects analyses^11^ will be reported separately and will assess whether the effect of PIN varies by clinic BPB status.

Analyses used R version 4.2.2 (R Foundation for Statistical Computing). Two-sided p < 0.05 was considered statistically significant.

### Results Participants

Across the 12 clinics, 2,303 patients had an abnormal FIT during the study period: 1,136 before and 1,167 after their clinic began the BPB (Figure 1). Of these, we randomly assigned 1,415 patients in the PIN trial: 698 to usual care and 717 to PIN; 888 were not randomly assigned to PIN or usual care, pending intervention design and deployment. We ascertained the 180-day outcome for all pre-BPB patients and 1,008 of 1,167 (86%) BPB-period patients; the remaining 159 had not reached 180 days of follow-up at the data cut-off. In the PIN trial, we ascertained the 180-day outcome for 668 of 698 usual-care and 679 of 717 PIN patients.

**Figure 1.**
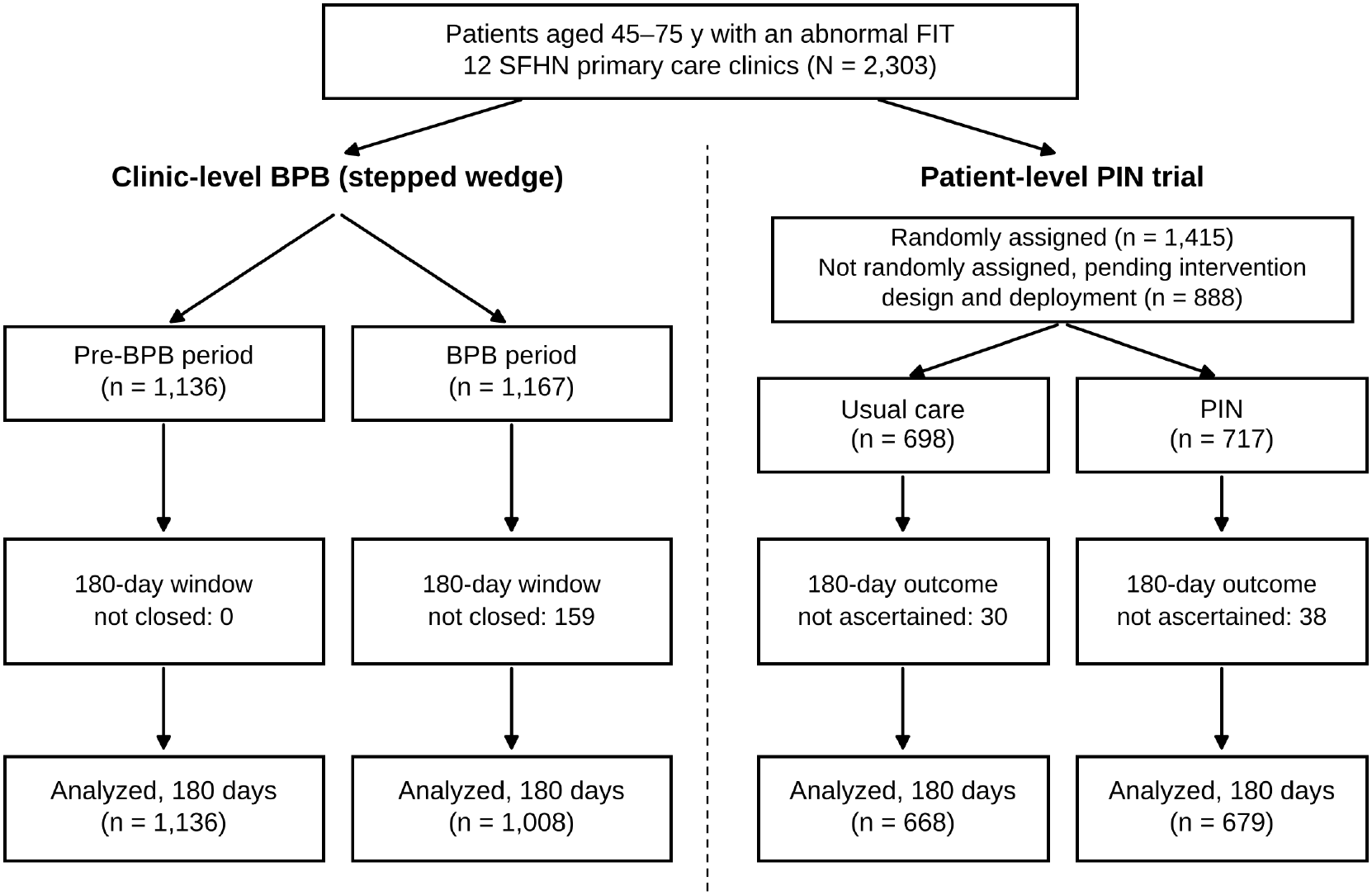
Participant flow. Patients with an abnormal FIT were classified by their clinic’s BPB period (left) and, for a subset, randomly assigned to PIN or usual care (right). The two designs ran concurrently in the same patients. The 180-day outcome was analyzed among patients whose 180-day window had closed by the data cut-off.

The population was diverse and largely low-income (Tables 1A and 1B). Median age was 60 years (IQR, 53–66); 52% preferred a language other than English, with 20% Cantonese and 19% Spanish; 48% had Medicaid; and 7.3% were homeless. Characteristics did not differ between the pre-BPB and BPB periods (all p ≥ 0.11). Baseline characteristics were generally similar between PIN and usual-care groups, although the PIN group included a somewhat lower proportion of women (43% vs 48%). Clinics differed substantially in patient mix (all p < 0.001; Supplementary Table S1). For example, 92% of patients at one clinic preferred a language other than English, and 33% of patients at another clinic were homeless. Race and ethnicity were unknown for 100 patients (4.3%); no other baseline variable was missing.

**Table A1.** Baseline characteristics of patients with an abnormal FIT, by clinic BPB period (stepped-wedge cohort)

| **Characteristic** | **Overall (N = 2,303)** | **Pre-BPB (N = 1,136)** | **BPB implemented (N = 1,167)** | **P-value** |
|---|---|---|---|---|
| Age at FIT, median (IQR), y | 60 (53–66) | 60 (54–66) | 60 (53–67) | 0.9 |
| Sex, No. (%) |  |  |  | 0.4 |
| Female | 1,034 (45) | 500 (44) | 534 (46) |  |
| Male | 1,269 (55) | 636 (56) | 633 (54) |  |
| Race and ethnicity, No. (%) |  |  |  | 0.2 |
| American Indian | 6 (0.3) | 3 (0.3) | 3 (0.3) |  |
| Asian | 846 (37) | 436 (38) | 410 (35) |  |
| Black or African American | 311 (14) | 155 (14) | 156 (13) |  |
| Hispanic/Latinx | 625 (27) | 288 (25) | 337 (29) |  |
| More than one race | 75 (3.3) | 44 (3.9) | 31 (2.7) |  |
| White | 340 (15) | 168 (15) | 172 (15) |  |
| Unknown | 100 (4.3) | 42 (3.7) | 58 (5.0) |  |
| Preferred language, No. (%) |  |  |  | 0.2 |
| English | 1,102 (48) | 541 (48) | 561 (48) |  |
| Cantonese | 464 (20) | 250 (22) | 214 (18) |  |
| Spanish | 448 (19) | 205 (18) | 243 (21) |  |
| Vietnamese | 61 (2.6) | 34 (3.0) | 27 (2.3) |  |
| Mandarin | 54 (2.3) | 28 (2.5) | 26 (2.2) |  |
| Russian | 45 (2.0) | 17 (1.5) | 28 (2.4) |  |
| Tagalog | 31 (1.3) | 17 (1.5) | 14 (1.2) |  |
| Toishanese | 25 (1.1) | 11 (1.0) | 14 (1.2) |  |
| Other | 73 (3.2) | 33 (2.9) | 40 (3.4) |  |
| Payor, No. (%) |  |  |  | 0.11 |
| Medicaid | 1,116 (48) | 573 (50) | 543 (47) |  |
| Medicare | 544 (24) | 250 (22) | 294 (25) |  |
| Healthy Workers | 523 (23) | 247 (22) | 276 (24) |  |
| Healthy SF | 94 (4.1) | 55 (4.8) | 39 (3.3) |  |
| Commercial | 13 (0.6) | 5 (0.4) | 8 (0.7) |  |
| Unknown | 13 (0.6) | 6 (0.5) | 7 (0.6) |  |
| Housing status, No. (%) |  |  |  | 0.9 |
| Housed | 2,136 (93) | 1,052 (93) | 1,084 (93) |  |
| Homeless | 167 (7.3) | 84 (7.4) | 83 (7.1) |  |

Abbreviations: BPB, Best Practice Bundle; FIT, fecal immunochemical test; IQR, interquartile range. Healthy SF is San Francisco's health access program for uninsured residents; Healthy Workers covers In-Home Supportive Services workers.

**Table 1B.**
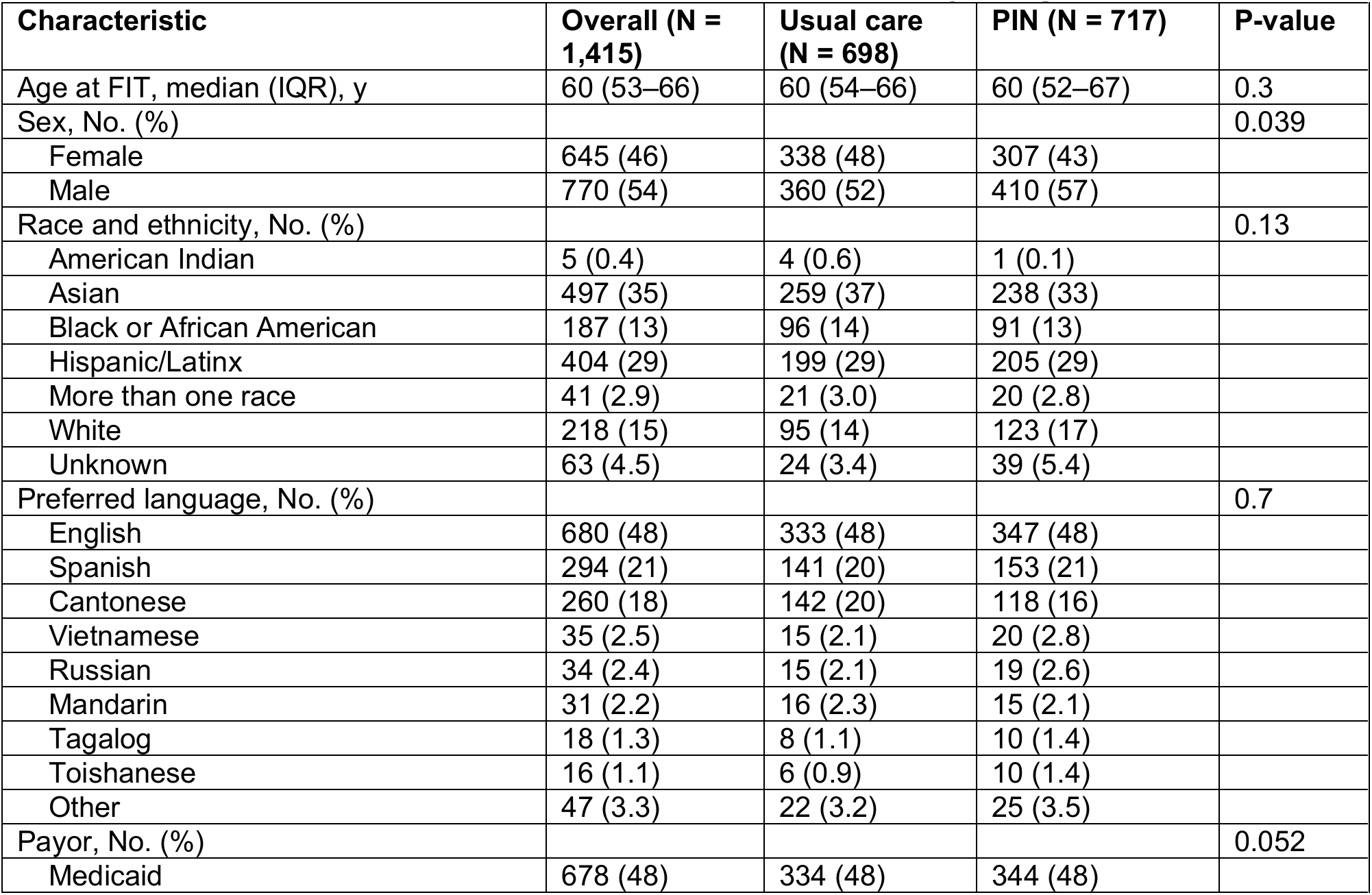
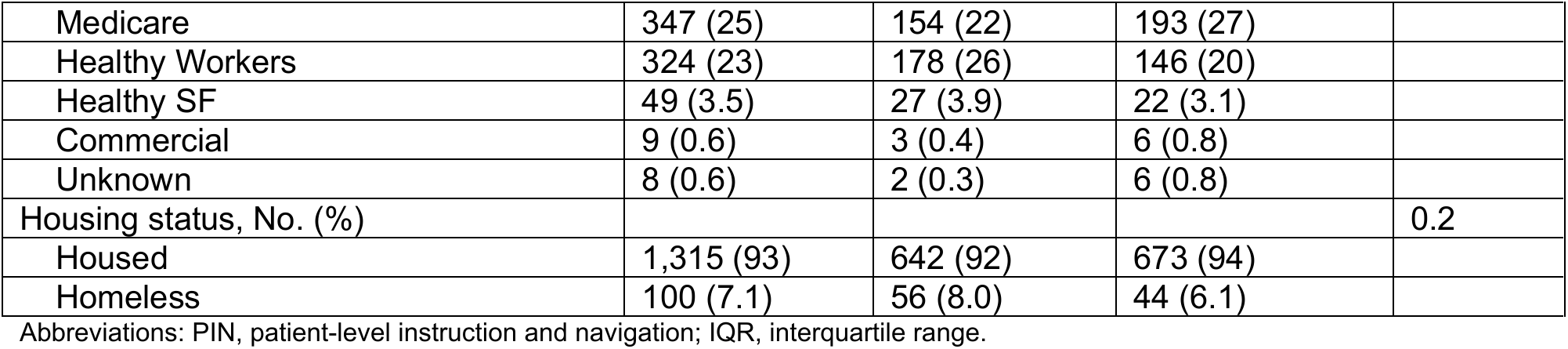
Baseline characteristics of patients in the PIN trial, by assignment.

| **Characteristic** | **Overall (N = 1,415)** | **Usual care (N = 698)** | **PIN (N = 717)** | **P-value** |
|---|---|---|---|---|
| Age at FIT, median (IQR), y | 60 (53–66) | 60 (54–66) | 60 (52–67) | 0.3 |
| Sex, No. (%) |  |  |  | 0.039 |
| Female | 645 (46) | 338 (48) | 307 (43) |  |
| Male | 770 (54) | 360 (52) | 410 (57) |  |
| Race and ethnicity, No. (%) |  |  |  | 0.13 |
| American Indian | 5 (0.4) | 4 (0.6) | 1 (0.1) |  |
| Asian | 497 (35) | 259 (37) | 238 (33) |  |
| Black or African American | 187 (13) | 96 (14) | 91 (13) |  |
| Hispanic/Latinx | 404 (29) | 199 (29) | 205 (29) |  |
| More than one race | 41 (2.9) | 21 (3.0) | 20 (2.8) |  |
| White | 218 (15) | 95 (14) | 123 (17) |  |
| Unknown | 63 (4.5) | 24 (3.4) | 39 (5.4) |  |
| Preferred language, No. (%) |  |  |  | 0.7 |
| English | 680 (48) | 333 (48) | 347 (48) |  |
| Spanish | 294 (21) | 141 (20) | 153 (21) |  |
| Cantonese | 260 (18) | 142 (20) | 118 (16) |  |
| Vietnamese | 35 (2.5) | 15 (2.1) | 20 (2.8) |  |
| Russian | 34 (2.4) | 15 (2.1) | 19 (2.6) |  |
| Mandarin | 31 (2.2) | 16 (2.3) | 15 (2.1) |  |
| Tagalog | 18 (1.3) | 8 (1.1) | 10 (1.4) |  |
| Toishanese | 16 (1.1) | 6 (0.9) | 10 (1.4) |  |
| Other | 47 (3.3) | 22 (3.2) | 25 (3.5) |  |
| Payor, No. (%) |  |  |  | 0.052 |
| Medicaid | 678 (48) | 334 (48) | 344 (48) |  |
| Medicare | 347 (25) | 154 (22) | 193 (27) |  |
| Healthy Workers | 324 (23) | 178 (26) | 146 (20) |  |
| Healthy SF | 49 (3.5) | 27 (3.9) | 22 (3.1) |  |
| Commercial | 9 (0.6) | 3 (0.4) | 6 (0.8) |  |
| Unknown | 8 (0.6) | 2 (0.3) | 6 (0.8) |  |
| Housing status, No. (%) |  |  |  | 0.2 |
| Housed | 1,315 (93) | 642 (92) | 673 (94) |  |
| Homeless | 100 (7.1) | 56 (8.0) | 44 (6.1) |  |

Abbreviations: PIN, patient-level instruction and navigation; IQR, interquartile range.

### Clinic-level Best Practice Bundle

Colonoscopy within 180 days was 44.1% in the pre-BPB period and 60.1% after BPB implementation (unadjusted difference of 16.0 percentage points; 95% CI, 11.8 to 20.1; p < 0.001; Table 2, Figure 2A). GI referral within 42 days was 69.8% before and 84.2% after BPB implementation (difference: 14.4 percentage points; 95% CI: 11.0 to 17.8). The proportion of patients referred at any time (91%) or scheduled at any time (78-79%) did not differ between periods. Among patients who completed colonoscopy, the median time from FIT to colonoscopy fell from 131 days (IQR, 82-251) to 86 days (IQR, 58-119; p < 0.001).

**Table 2.** Outcomes before and after clinic implementation of the Best Practice Bundle (stepped-wedge cohort, N = 2,**303)**

| Outcome | Pre-BPB, n/N (%) | BPB implemented, n/N (%) | Risk difference, percentage points (95% CI) | P-value |
|---|---|---|---|---|
| **Primary** |  |  |  |  |
| Colonoscopy within 180 days (a) | 501/1,136 (44.1) | 606/1,008 (60.1) | 16.0 (11.8 to 20.1) | <0.001 |
| **Secondary** |  |  |  |  |
| GI referral within 42 days (b) | 793/1,136 (69.8) | 974/1,157 (84.2) | 14.4 (11.0 to 17.8) | <0.001 |
| Days from FIT to colonoscopy, median (IQR) (c) | 131 (82–251) | 86 (58–119) |  | <0.001 |
| Days from FIT to GI referral, median (IQR) (c) | 7 (4–21) | 6 (3–12) |  | <0.001 |
| **Supporting measures** |  |  |  |  |
| GI referral at any time | 1,036/1,136 (91.2) | 1,067/1,167 (91.4) | 0.2 (−2.1 to 2.6) | >0.9 |
| Colonoscopy scheduled at any time | 901/1,136 (79.3) | 911/1,167 (78.1) | −1.2 (−4.6 to 2.1) | 0.5 |
| Colonoscopy at any time by data cut-off (d) | 765/1,136 (67.3) | 690/1,167 (59.1) |  | <0.001 |

All estimates unadjusted. Risk differences with 95% CIs; p-values from the chi-square or Wilcoxon rank-sum test. (a) Excludes 159 BPB-period patients whose 180-day window had not closed. (b) Excludes 10 BPB-period patients whose 42-day window had not closed. Scheduling within 56 days is not reported because scheduling dates were not captured accurately before the BPB. (c) Among patients who reached the step, with values capped at 180 days, days to colonoscopy were 131 (82–180) vs 86 (58–119). (d) Not comparable between periods because pre-BPB patients had longer follow-up.

**Figure 2.**
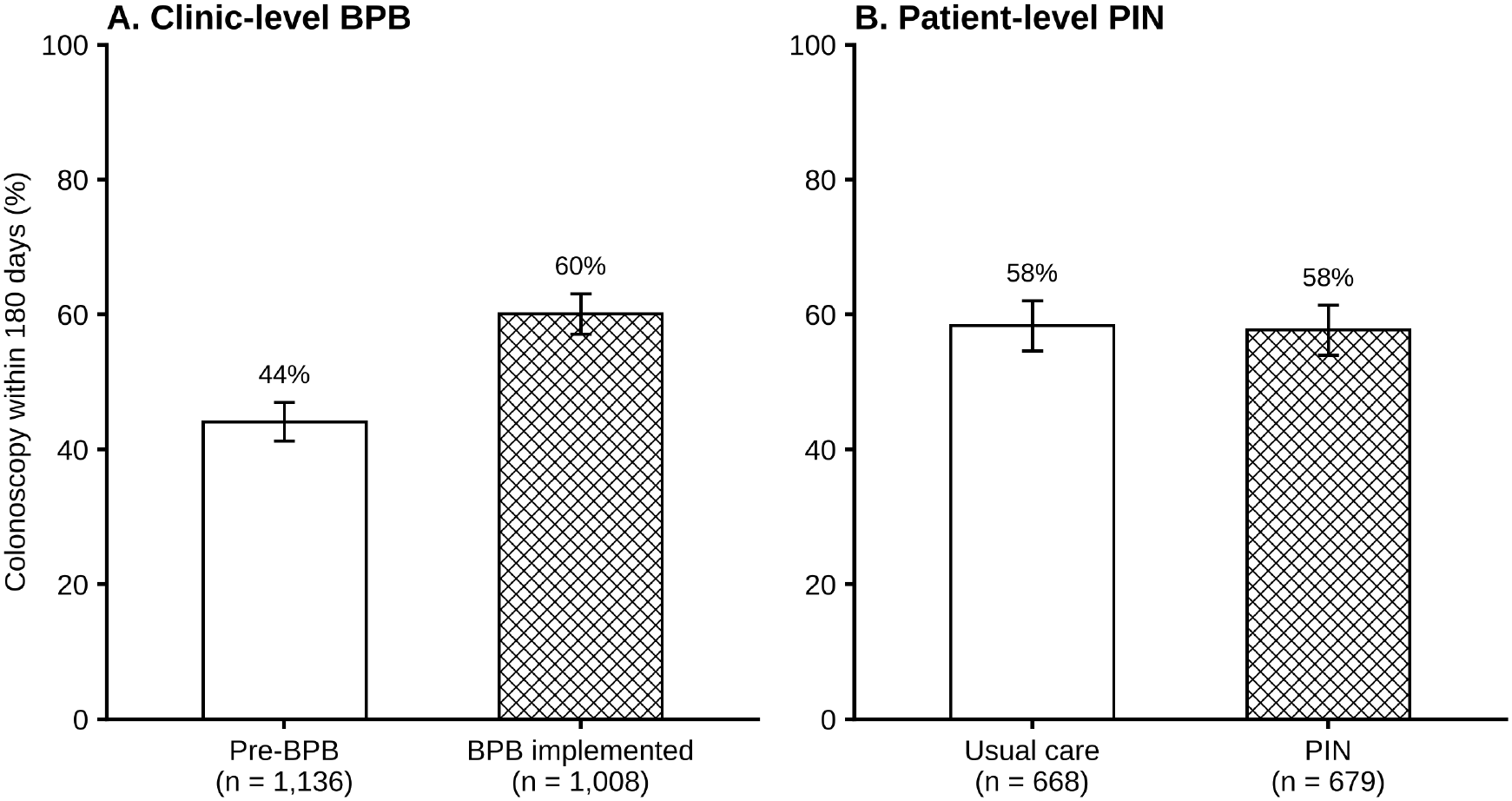
Colonoscopy within 180 days of the abnormal FIT. (A) Before and after the clinic-level Best Practice Bundle (pre-BPB, 501/1,136; BPB implemented, 606/1,008). (B) By PIN assignment (usual care, 390/668; PIN, 392/679). Denominators exclude patients whose 180-day window had not yet closed or whose outcomes were not ascertained. Error bars, 95% CI, unadjusted.

### Patient-level PIN trial

Colonoscopy within 180 days did not differ between arms: 58.4% with usual care and 57.7% with PIN (difference, -0.7 points; 95% CI, -5.9 to 4.6; p = 0.9; Table 3, Figure 2B). Among patients enrolled before BPB implementation during January to October 2024, colonoscopy completion within 180 days was 56.6% with PIN (47/83) and 50.6% with usual care (39/77), a difference of 6.0 percentage points (95% CI –9.5% to 21.4%; p = 0.4). PIN also did not significantly improve GI referral within 42 days (84.2% vs 80.3%; p = 0.06) or scheduling within 56 days (46.7% vs 43.8%; p = 0.3).

**Table 3.** Outcomes by PIN assignment (N = 1,415)

| Outcome | Usual care, n/N (%) | PIN, n/N (%) | Risk difference, percentage points (95% CI) | P-value |
|---|---|---|---|---|
| **Primary** |  |  |  |  |
| Colonoscopy within 180 days (a) | 390/668 (58.4) | 392/679 (57.7) | −0.7 (−5.9 to 4.6) | 0.9 |
| **Secondary** |  |  |  |  |
| GI referral within 42 days | 588/698 (84.2) | 576/717 (80.3) | −3.9 (−7.9 to 0.1) | 0.064 |
| Colonoscopy scheduled within 56 days | 326/698 (46.7) | 314/717 (43.8) | −2.9 (−8.1 to 2.3) | 0.3 |
| Days from FIT to colonoscopy, median (IQR) (b) | 84 (56–122) | 88 (58–131) |  | 0.3 |
| Days from FIT to GI referral, median (IQR) (b) | 6 (3–11) | 6 (3–14) |  | 0.4 |
| Days from FIT to scheduling, median (IQR) (b) | 39 (26–77) | 38 (26–82) |  | 0.8 |
| **Supporting measures** |  |  |  |  |
| GI referral at any time | 653/698 (93.6) | 662/717 (92.3) | −1.2 (−3.9 to 1.5) | 0.4 |
| Colonoscopy scheduled at any time | 582/698 (83.4) | 575/717 (80.2) | −3.2 (−7.2 to 0.8) | 0.14 |
| Colonoscopy at any time by data cut-off | 459/698 (65.8) | 467/717 (65.1) |  | 0.8 |

All estimates unadjusted. (a) Excludes 30 usual-care and 38 PIN patients without 180-day ascertainment. (b) Among patients who reached the step, values capped at 180 days were identical.

### Text-message engagement

Of 313 PIN patients with a scheduled colonoscopy who received a text reminder, 180 (57.5%; 95% CI, 52.0–62.9) replied (Supplementary Figure S1). Of those who replied, 150 (83%) confirmed, 16 (8.9%) rescheduled, and 14 (7.8%) canceled; 161 (89.4%) completed colonoscopy. Of 96 patients who did not reply and received a reminder call, 34 (35%) confirmed, 45 (47%) received a voicemail, and 14 (15%) could not be reached; 79 (82.3%) completed colonoscopy. Due to operational oversight, 37 patients who did not reply did not receive a reminder call; 29 (78%) completed colonoscopy. These are descriptive results among patients who had already scheduled a colonoscopy, not comparisons with usual care.

## Discussion

In this preliminary analysis of a multilevel intervention to improve diagnostic colonoscopy after abnormal FIT in a diverse safety-net population, implementation of the clinic-level BPB was associated with substantial improvements in timely follow-up. Referral rates and colonoscopy completion within 180 days substantially increased after implementation of the BPB. Among patients who completed colonoscopy, the median time from an abnormal FIT to colonoscopy decreased substantially. In contrast, the randomized patient-level PIN intervention did not improve colonoscopy completion within 180 days, timely referral, or scheduling compared with usual care.

The improvement observed after BPB implementation supports the importance of addressing clinic-level processes in the multistep pathway from abnormal FIT to diagnostic colonoscopy.^8^ Prior studies have identified provider reminders, automated referrals, patient tracking, and audit and feedback as strategies that may improve follow-up after abnormal cancer screening.^4^ Our prior work in SFHN similarly identified inconsistent use of registries, variable communication practices, and lack of standardized workflows as potential contributors to incomplete follow-up.^12^ We designed the BPB to address these gaps through systematic patient tracking, standardized EHR documentation and communication, and audit and feedback. Notably, the proportion of patients ever referred or ever scheduled changed little after BPB implementation, whereas the rates of timely referral and 180-day colonoscopy completion increased. This pattern suggests that strengthening clinic workflows may improve the timeliness and reliability of existing care processes rather than simply increasing the likelihood that referral or scheduling eventually occurs.

The lack of observed benefit from PIN contrasts with prior evidence that patient navigation and reminders can facilitate diagnostic follow-up.^4,13^ Several factors may help explain this finding. One possible explanation is that colonoscopy after an abnormal FIT requires multiple transitions among primary care, gastroenterology, and patients,^8,9^ such that patient-facing support alone may be insufficient to overcome barriers elsewhere in the care pathway. We designed PIN as a technology-supported approach that reserved staff for low-touch prompts and reminders to clinical care teams, rather than as a traditional intensive navigation model. Colonoscopy completion was high among patients who engaged with the intervention, with most respondents confirming their appointments. Further analyses of intervention reach, engagement, patient subgroups, and reasons for incomplete follow-up may help identify whether particular patients or stages of the care pathway benefited from PIN despite the absence of an overall effect.

These findings also underscore the value of examining multilevel interventions within the context in which they are implemented.^8^ SFHN serves a heterogeneous safety-net population, and clinics differed substantially in patient characteristics. Implementation of the BPB may likewise have varied across clinics. Planned implementation and qualitative analyses will provide additional information about intervention reach, acceptability, feasibility, local workflow adaptation, and barriers and facilitators to implementation and may help explain variation in quantitative outcomes.

Several limitations should be considered. First, these are preliminary unadjusted analyses, though randomization was fairly balanced in BPB and PIN subgroups. The BPB comparison is based on pre- and post-implementation periods in a stepped-wedge rollout in which the timing of clinic implementation was not randomized; therefore, secular trends and other contemporaneous changes may contribute to the observed differences, and the findings should not be interpreted as causal estimates of BPB effectiveness. Prespecified mixed-effects analyses will account for calendar time, clinic-level clustering, patient characteristics, and concurrent PIN assignment. Second, 159 BPB-period patients had not yet reached 180 days of follow-up at the data cutoff and were excluded from the 180-day analysis, whereas all pre-BPB patients had completed the follow-up window. Third, exposure to individual PIN components depended on where patients encountered barriers along the follow-up pathway; therefore, assignment to PIN did not necessarily result in receipt of all intervention components. Finally, we conducted the study within a single integrated urban safety-net health system, which may limit generalizability to other settings.

Preliminary findings suggest that implementation of clinic-level systems for tracking and managing abnormal FIT follow-up was associated with more timely diagnostic colonoscopy. In contrast, the randomized technology-supported PIN intervention did not improve colonoscopy completion. These findings highlight the importance of addressing the multiple care transitions and organizational processes involved in abnormal FIT follow-up. Completion of the prespecified effectiveness analyses and integration of implementation and qualitative findings will provide a more comprehensive understanding of which intervention components work, for whom, and under what conditions.

## Data Availability

The data underlying this study are not currently publicly available. Data sharing will be considered following publication of the study findings, subject to applicable institutional, ethical, and data-use requirements.

## Declarations

### Ethics approval and consent to participate

The UCSF Institutional Review Board (IRB #17-23991) approved the study with a waiver of informed consent.

### Consent for publication

Not applicable.

