## Supplementary Figure S1 for "Multilevel Interventions to Improve Colonoscopy Completion After Abnormal Fecal Immunochemical Testing in a Safety-Net Health System: Preliminary Findings from the IMPACTT Study"

### Supplementary Material

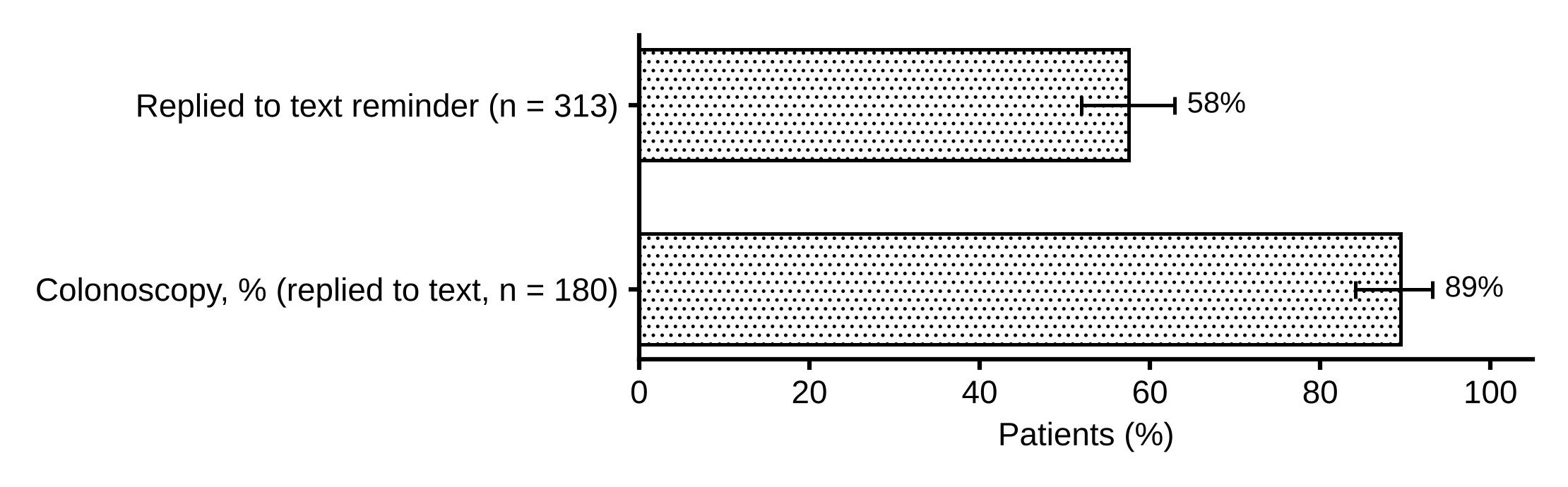

**Supplementary Figure S1. Text message response and colonoscopy completion among PIN patients with a scheduled colonoscopy.** Replied to text reminder, 180/313; colonoscopy among patients who replied, 161/180. Error bars, 95% CI.
