## Supplementary Table S1 for "Multilevel Interventions to Improve Colonoscopy Completion After Abnormal Fecal Immunochemical Testing in a Safety-Net Health System: Preliminary Findings from the IMPACTT Study"

### Supplementary Material

**Supplementary Table S1. Patient characteristics by clinic (N = 2,303), sorted by clinic size**

| **Clinic** | **N** | **Median age, y** | **Non-English language, %** | **Homeless, %** | **Medicaid, %** | **Female, %** | **Asian, %** | **Black, %** | **Hispanic, %** | **White, %** |
| --- | --- | --- | --- | --- | --- | --- | --- | --- | --- | --- |
| Clinic 1 | 448 | 58 | 60 | 4 | 56 | 54 | 25 | 11 | 46 | 11 |
| Clinic 2 | 400 | 60 | 50 | 8 | 56 | 46 | 34 | 10 | 36 | 14 |
| Clinic 3 | 259 | 62 | 92 | 2 | 21 | 54 | 95 | 1 | 1 | 2 |
| Clinic 4 | 205 | 60 | 69 | 1 | 39 | 41 | 55 | 6 | 28 | 3 |
| Clinic 5 | 180 | 60 | 17 | 33 | 61 | 36 | 12 | 22 | 22 | 31 |
| Clinic 6 | 174 | 61 | 66 | 0 | 29 | 53 | 64 | 1 | 2 | 23 |
| Clinic 7 | 152 | 60 | 23 | 3 | 57 | 42 | 11 | 51 | 22 | 7 |
| Clinic 8 | 124 | 60 | 44 | 6 | 62 | 41 | 10 | 11 | 45 | 23 |
| Clinic 9 | 114 | 62 | 43 | 1 | 46 | 48 | 30 | 28 | 18 | 19 |
| Clinic 10 | 108 | 66 | 30 | 27 | 53 | 26 | 26 | 23 | 18 | 24 |
| Clinic 11 | 70 | 62 | 34 | 3 | 54 | 37 | 21 | 24 | 30 | 14 |
| Clinic 12 | 69 | 58 | 17 | 14 | 51 | 6 | 4 | 4 | 39 | 45 |

All between-clinic comparisons, p < 0.001.
